# Optimal annual hospital volume for open surgical repairs for abdominal aortic aneurysms: a protocol of systematic review and dose-response meta-analysis

**DOI:** 10.1101/2025.08.18.25333932

**Authors:** Eiki Kanemaru, Kanako Sasaki, Tomoaki Miyake, Hisashi Noma, Takahisa Goto, Takahiro Mihara

**Affiliations:** Department of Health Data Science, Graduate School of Data Science, Yokohama City University; Department of Anesthesiology, Yokohama City University Hospital; Department of Interdisciplinary Statistical Mathematics, The Institute of Statistical Mathematics; Department of Anesthesiology, Yokohama City University School of Medicine

## Abstract

**Introduction:** Centralization of open surgical repairs for abdominal aortic aneurysms (AAA) has been advocated to improve patient outcomes and reduce healthcare costs. To promote centralization, it is essential to establish an evidence-based optimal threshold for annual number of procedures performed at each hospital. This review aims to conduct a dose-response meta-analysis to determine the optimal annual hospital volume for open AAA repairs.

**Methods and analysis:** This protocol was developed in accordance with the 2015 Preferred Reporting Items for Systematic Reviews and Meta-Analyses Protocols (PRISMA-P) guidelines. We will include all prospective or retrospective cohort studies evaluating a volume-outcome relationship in open AAA repairs, provided they report at least two distinct hospital volumes. Studies reporting only a single hospital volume or focusing solely on surgeon-specific volumes will be excluded. Relevant studies will be searched in PubMed, EMBASE, Cochrane Library, and Web of Science from inception to 13 August 2025, without language restrictions. Three authors will independently screen and select eligible studies, extract relevant data, and assess the risk of bias. The primary outcome is short-term mortality, defined as in-hospital and/or 30-day mortality, analyzed in relation to annual hospital volume through both dose-response and pairwise meta-analyses. A dose-response meta-analysis will be performed for studies reporting three or more categories of annual hospital volume. For pairwise meta-analysis, studies with two comparison groups will be analyzed directly, whereas studies with three or more comparison groups will be converted into binary comparisons. The certainty of evidence for each outcome will be assessed using the Grading of Recommendations Assessment, Development and Evaluation (GRADE) approach and the Core GRADE approach.

**Ethics and dissemination:** As this study will be based solely on previously published data, ethics approval is not required. The findings will be disseminated through publication in a peer-reviewed journal.

**PROSPERO registration number:** CRD420251121357

## INTRODUCTION

### Rationale

Centralizing health care services is one of the most important contemporary themes in health care policy and may be particularly important for high-risk surgeries (1). Recently, centralization efforts have been made for various clinical procedures, such as oncological and coronary interventions (2, 3). Within this framework, high-risk procedures should be concentrated in centers of specialized expertise, where annual case volume is the principal marker of proficiency. Both historical and contemporary studies have consistently demonstrated the relationship between volume and outcome in various types of surgeries (4, 5).

Various health care settings and organizations have confirmed the established volume-outcome relationship for open surgical repairs for abdominal aortic aneurysms (AAA). Large vascular surgical registries from 11 countries showed that the highest volume centers significantly reduced post-operative mortality rate compared with the lowest volume centers in unruptured AAA repairs (6, 7). The association between volume and outcome has also been demonstrated in ruptured AAA repairs. In a meta-analysis including 13 studies with a total of 120,116 patients, patients treated in low-volume hospitals had a statistically significantly higher post-operative mortality rate than those treated in high-volume hospitals (8).

Based on the current evidence of a volume-outcome relationship in open surgical repairs for AAA, it is justifiable to recommend a minimum surgical volume for aortic centers. European Society for Vascular Surgery recommends that centers performing open AAA repairs should not have an annual caseload of less than 15 (9). The Society for Vascular Surgery guidelines endorse at least 10 open AAA repairs per year as a minimum aortic practice requirement (10). However, the annual case volume thresholds recommended in these guidelines are based primarily on individual dose-response studies, which vary considerably in sample size and methodology. In addition, previous studies were confined to the categorical assessment of case volume, resulting in the possibility that a potential non-linear volume-outcome relation might be missed. Therefore, this study aims to conduct a dose-response meta-analysis to establish more robust evidence for the optimal annual hospital volume for open AAA repairs.

### Objectives

The objective of this systematic review and dose-response meta-analysis is to investigate the relationship between annual hospital volume and the mortality of patients who undergo open AAA repairs.

## METHODS

### Study design

The objectives and methodology of this systematic review were pre-specified in a protocol registered with PROSPERO (International Prospective Register of Systematic Reviews in Health and Social Care) under the registration number CRD420251121357. The reporting of this review will adhere to the Preferred Reporting Items for Systematic Reviews and Meta-Analyses (PRISMA) guidelines (11).

### Eligibility criteria

We will include all published prospective or retrospective cohort studies assessing the association between annual hospital volume and the mortality of patients, who are 18 years old and above, undergoing open AAA repairs. Studies are needed to report on (1) the annual case volume of open AAA repairs and (2) the postoperative mortality (in-hospital and/or 30-day mortality). The studies that report at least two different hospital volumes will be included. If the studies from the same center or database reported on separate cohorts in different periods, they will be included and assessed separately. The conference abstracts will be eligible for publication to mitigate the risk of publication bias. We will exclude the studies that don’t report on the annual hospital volume of open AAA repairs and the postoperative mortality, those that report data on the single hospital volume, and those that report surgeon volume. To avoid double-counting cohorts derived from the same underlying database (e.g., national registries or claims data), when two or more studies appear to use overlapping data, we will include only the study with the largest eligible sample size and exclude the others.

### Information sources

PubMed, EMBASE, Cochrane Library, and Web of Science will be systematically searched for eligible articles. Furthermore, the reference lists of included articles will be screened for potential missed articles. The search will be initiated from the databases’ inception, and the last search will be performed on 13 August 2025 for all databases (E.K., K.S., and T. Miyake).

### Search strategy

Search strategies will be developed using medical subject headings (MeSH) and free text words relating to AAA surgery, hospital volume, and treatment outcome. The proposed search strategy for PubMed is presented in Table 1. The search strategies for other databases are available on request.

**Table 1.** Search strategy for PubMed.

| Number | Query |
| --- | --- |
| #1 | "aortic aneurysm, abdominal/surgery"[MeSH Terms] |
| #2 | "abdominal aortic aneurysm*"[Title/Abstract] |
| #3 | "Aortic Surgery"[Title/Abstract] |
| #4 | "aortic aneurysm"[Title/Abstract] |
| #5 | "AAA"[Title/Abstract] |
| #6 | "Cardiovascular Surgery"[Title/Abstract] |
| #7 | "Acute Aortic Syndrome"[MeSH Terms] |
| #8 | "dissecting aneurysm"[Title/Abstract] |
| #9 | #1 OR #2 OR #3 OR #4 OR #5 OR #6 OR #7 OR #8 |
| #10 | "surgeons/statistics and numerical data"[MeSH Terms] |
| #11 | "hospitals, high-volume"[MeSH Terms] |
| #12 | "hospitals, low-volume"[MeSH Terms] |
| #13 | "high volume"[Title/Abstract] |
| #14 | "low volume"[Title/Abstract] |
| #15 | "large volume"[Title/Abstract] |
| #16 | "small volume"[Title/Abstract] |
| #17 | "case volume"[Title/Abstract] |
| #18 | "surgical volume"[Title/Abstract] |
| #19 | "hospital volume"[Title/Abstract] |
| #20 | "center volume"[Title/Abstract] |
| #21 | "institutional volume"[Title/Abstract] |
| #22 | "procedural volume"[Title/Abstract] |
| #23 | "case load"[Title/Abstract] |
| #24 | "caseload"[Title/Abstract] |
| #25 | "professional competence"[Title/Abstract] |
| #26 | "expertise"[Title/Abstract] |
| #27 | "provider volume"[Title/Abstract] |
| #28 | "volume-outcome"[Title/Abstract] |
| #29 | #10 OR #11 OR #12 OR #13 OR #14 OR #15 OR #16 OR #17 OR #18 OR #19 OR #20 OR #21 OR #22 OR #23 OR #24 OR #25 OR #26 OR #27 OR #28 |
| #30 | "Treatment Outcome"[MeSH Terms] |
| #31 | "Patient Outcome Assessment"[MeSH Terms] |
| #32 | "Postoperative Complications"[MeSH Terms] |
| #33 | "Mortality"[MeSH Terms] |
| #34 | "Hospital Mortality"[MeSH Terms] |
| #35 | "Survival Rate"[MeSH Terms] |
| #36 | "Length of Stay"[MeSH Terms] |
| #37 | "Mortality"[Title/Abstract] |
| #38 | "survival"[Title/Abstract] |
| #39 | "prognosis"[Title/Abstract] |
| #40 | "Treatment Outcome"[Title/Abstract] |
| #41 | "morbidity"[MeSH Terms] |
| #42 | "Length of Stay"[Title/Abstract] |
| #43 | #30 OR #31 OR #32 OR #33 OR #34 OR #35 OR #36 OR #37 OR #38 OR #39 OR #40 OR #41 OR #42 |
| #44 | #9 AND #29 AND #43 |

### Selection process

Three authors (E.K., K.S., and T. Miyake) will independently perform the selection process. Studies will be screened for eligibility based on title and abstract. We will use the web-based application Rayyan (http://www.rayyan.ai) for this phase, after which eligible full-texts will be evaluated for final inclusion. Any potential disagreement will be resolved through discussion.

### Data collection process and data items

Data collection will be performed independently by three authors (E.K., K.S., and T. Miyake). Discrepancies will be resolved via discussion including a review author (T. Mihara) with expertise in systematic review and meta-analysis if necessary. We will contact the study authors for clarification if data are missing or presented ambiguously. A predefined data extraction sheet will be developed to collect the most relevant study characteristics, including study title, year of publication, authors, journal, country, data source, number of years when patients were recruited, diagnosis (unruptured or ruptured), urgency (elective or urgent), aneurysm characteristics (e.g. complex aneurysms requiring suprarenal aortic clamp, interposition or bypass graft to any visceral branch, or surgery for infected aneurysms), annual hospital volume, the total number of patients and the number of deaths for each annual hospital volume category, in-hospital and/or 30-day mortality, 1-year mortality, length of hospital stay, effect measures, and adjusted covariates including age, sex, disease severity, comorbidity, and hospital and regional characteristics.

### Outcomes and effect measures

The primary outcome is short-term mortality, defined as in-hospital and/or 30-day mortality, in relation to annual hospital volume, as assessed by dose-response meta-analysis and pairwise meta-analysis. The secondary outcomes are 1-year mortality and length of hospital stay. The mortality will be measured as the risk, odds, or hazard ratios calculated using the number of events with the lowest annual hospital volume as the reference category. The length of hospital stay will be measured in days.

### Risk of bias assessment

The Newcastle-Ottawa scale (NOS) will be used to assess study quality objectively. The NOS comprises Selection (representativeness of the exposed cohort, selection of the non-exposed cohort, ascertainment of exposure, demonstration that outcome of interest was not present at start of study), Comparability (comparability of cohorts on the basis of the design or analysis controlled for confounders), and Outcome (assessment of outcome, long enough follow-up for outcomes to occur, adequacy of follow-up of cohorts) (12). The studies will be classified as high (7–9) or low quality (0–6). The quality assessment will be performed by three authors (E.K., K.S., and T. Miyake).

### Data synthesis

#### 1) Dose-response meta-analysis

A dose-response meta-analysis will be performed for studies reporting three or more categories of annual hospital volume. We will treat volume as a continuous exposure by assigning a representative value to each category. Study-specific log–relative risks (the risk, odds, or hazard ratios) and their within-study variance–covariance matrices (to account for the shared reference group) will be estimated using the Greenland & Longnecker generalized least squares trend method, as implemented in the R package *dosresmeta*. In addition, to maximize the use of all available data, we will also perform analyses that don’t adjust for within-study correlations for studies in which the number of cases or participants is not reported. The estimates will be pooled using a multivariate random-effects model. To assess non-linearity, we will fit restricted cubic spline models and compare them with the linear model using a Wald test of the spline terms. If the non-linearity test is not significant, we will report the pooled linear trend; otherwise, we will present the spline curve and predicted risks at clinically relevant volumes (13). The elbow method will be used to determine the optimal annual hospital volume (5).

#### 2) Pairwise meta-analysis

A pairwise meta-analysis will be performed for studies that report two comparison groups. Studies that report three or more comparison groups will also be included in a pairwise meta-analysis by converting them to binary comparisons using the lowest and highest annual hospital volume groups. The risk, odds, or hazard ratios (for dichotomous outcomes) and mean differences (for continuous outcomes) will be summarized with 95% confidence intervals. A random-effects model will be applied to aggregate the results. Statistical heterogeneity will be assessed using the I^2^ statistic, with I^2^ > 50% indicating substantial heterogeneity. Subgroup analysis will be conducted to explore potential causes for studies showing high heterogeneity.

### Analysis of subgroups or subsets

To explore potential causes of heterogeneity, subgroup analyses will be performed based on diagnosis (unruptured or ruptured), procedural urgency (elective or urgent), geographic location, study period (using 2005 as a cutoff to reflect the transition before and after the prevalence of endovascular treatment), and volume grouping (dichotomies, tertiles, quartiles, quintiles, sextiles, or Nonilies). To ensure the robustness of the findings, sensitivity analyses will be performed by integrating studies based on the most frequently reported outcome measure (e.g., the risk, odds, or hazard ratios), integrating studies with outcome adjusted for covariables, excluding studies considered to have a potential risk of bias (NOS < 7), excluding studies targeting complex aneurysms, and excluding studies that used failure to rescue as the study endpoint.

### Publication bias assessment and adjustment

Publication bias or small-study effects will be evaluated using funnel plots and Egger’s regression asymmetry test. A p-value < 0.10 in Egger’s test will be interpreted as evidence of asymmetry.

### Evaluation of the certainty of evidence

We will use the Grading of Recommendations Assessment, Development and Evaluation (GRADE) approach and the Core GRADE approach to assess the certainty of evidence for each outcome (14–23). The certainty of evidence will be evaluated across the following domains: risk of bias, imprecision, indirectness, inconsistency, publication bias, dose-response, and large effect. When deciding whether to rate down certainty for imprecision, we will consider whether the confidence interval crosses the chosen threshold, which is the minimal important difference (MID) (18). We will judge the magnitude of inconsistency by evaluating the extent to which point estimates differ and the degree to which confidence intervals overlap (19). When deciding whether to rate down certainty for inconsistency, we will evaluate where individual study estimates lie in relation to the MID (19). The MID for mortality is defined as 0.9 of risk, odds, or hazard ratio, using the category with the smallest number of cases as the reference. The MID for the length of hospital stay is defined as a 1-day decrease. The certainty of evidence will be categorized as high, moderate, low, and very low.

## LIMITATIONS AND IMPLICATIONS

There are some limitations. First, as the primary outcomes are reported as various effect measures, such as the risk, odds, or hazard ratios, the pooled estimate derived from all types of effect measures should be interpreted cautiously. To address the first limitation, we plan to conduct a sensitivity analysis by restricting the analysis to the most commonly used effect measure to evaluate whether the results remain consistent. If the findings are similar, this would support the robustness of the results. However, if substantial discrepancies exist, we will interpret the results analyzed by the single most frequently reported outcome measure. Second, as the primary outcomes comprise both covariate-adjusted and unadjusted effect estimates, the pooled estimate should be interpreted cautiously. To address the second limitation, a sensitivity analysis that includes only studies with covariate-adjusted effect estimates will be conducted to evaluate the robustness of the findings.

Centralization of open AAA repairs has been advocated to improve patient outcomes and reduce healthcare costs. To promote centralization, it is essential to establish an evidence-based optimal threshold for annual number of procedures performed at each hospital. However, previous studies investigating the volume-outcome relationship have primarily relied on categorical assessments of case volume, which may have overlooked potential non-linear volume-outcome associations. The findings from this dose-response meta-analysis will indicate more robust evidence for the optimal annual hospital volume for open AAA repairs. If the dose-response relationship proves to be linear, it would support the findings of previous studies. Conversely, if a non-linear association is observed, this review may provide a more accurate estimate of the appropriate annual case volume threshold.

## ETHICS AND DISSEMINATION

As this study will be based solely on previously published data, ethics approval is not required. The results will be submitted to a peer-reviewed journal for publication. The results will also be presented at relevant conferences. Any significant amendments to the study protocol will be clearly documented, along with an explanation of the changes, their justification, and the date they are made.

## Data availability statement

Since this protocol did not involve generating or analyzing any datasets, data sharing is not applicable.

## Authors’ contributions

E.K. and T. Mihara designed the review. K.S. and T. Miyake helped develop the review protocol. E.K. and T. Mihara wrote the draft. T. Mihara and H. Noma supervised all parts of the review protocol. All authors contributed to the article and approved the submitted version.

